# A Qualitative Study Regarding Messages of the COVID-19 Vaccine from Vaccinated Healthcare Providers and Healthy Adults

**DOI:** 10.1101/2022.03.24.22272878

**Authors:** Shuji Sano, Satomi Sato, Norio Ohmagari, Osamu Takahashi

## Abstract

1.

**Background:** To promote the vaccination against COVID-19, person-to-person communication from vaccinated people will play an important role. The objectives of this study are to identify what messages were shared by vaccinated people, and the relationship between these messages and their background.

**Methods:** This study was an exploratory and prospective basis with individual interviews. The participants were healthcare providers and healthy adults who were recruited at a vaccination site in Chuo-City, Tokyo. The online interviews were conducted using a semi-structured interview. Based on the Health Belief Model (HBM), the participants were asked about their perspectives on vaccines and what they talked about after their vaccination. The interviews were categorized into each item of the HBM and analyzed using NVivo software.

**Results:** During August to October 2021, five healthcare providers and seven healthy adults were enrolled in the study. One healthy adult could not be contacted resulting in a total of 11 participants interviewed. Both the healthcare providers and the healthy adults mainly talked about side effects after their vaccination, and to ease the other persons’ concerns based on their experience. Meanwhile, there were differences in the recommendations for vaccination between the two groups. The healthcare providers were strongly aware of the severity of COVID-19 infection and recommended vaccination to others as a useful measure to suppress becoming severely ill. On the other hand, the healthy adults recommended the vaccine with varying degree depending on their expectations and concerns about the vaccine and external factors such as living with a family member.

**Conclusion:** Both the healthcare providers and healthy adults shared similar messages to ease the vaccination concerns of others. However, their vaccine recommendation level was varied, which may be influenced not only by expectations and concerns toward the vaccine, but also by external factors such as family members living together.

## 2. INTRODUCTION

### 2.1. Background

The spread of the COVID-19 has threatened and changed our daily lives. In such a situation, vaccination plays an important role in infection control by promoting herd immunity [1]. The Japanese government has promoted COVID-19 vaccination for the entire population, and those who wish to be vaccinated can do so for free [2].

Currently, the mRNA vaccines developed by Pfizer (Comirnaty™ intramuscular injection), and Moderna (Spikevax™ intramuscular injection), and the adenovirus vector vaccine developed by AstraZeneca (Vaxzevria™ intramuscular injection) have been approved by the Ministry of Health, Labour and Welfare in Japan [2], and have been available for clinical use. However, some people were hesitant or unwilling to receive the COVID-19 vaccine due to a lack of long-term data and concerns about the safety of the vaccine influenced by information being spread by mass media or social network services [3,4].

In order to promote the vaccination, the Japanese government has provided the public with information through websites and TV commercials to aid decision-making on whether to vaccinate based on a comprehensive consideration of risks and benefits [2]. The reliability of information sources is a very important aspect; information from the Internet have been unreliable due to insufficient or incorrect information [5], and have influenced some people to not trust the information from the government or media, particularly those hesitant to receive the COVID-19 vaccine [6]. On the other hand, a previous report explained that person-to-person communication was more effective in influencing health behaviors [7]. In addition, information from family members and close friends are very trustworthy because of their strong relationship [6,8]. This can be explained by the interpersonal communication theory. This theory is based on human relationships from which health behaviors emerge, thereby affecting a variety of health outcomes[9,10]. Therefore, person-to-person communication, especially among close persons, have a high impact on health behaviors.

Based on this, our assumption is that person-to-person communication with those who had been vaccinated for COVID-19 was considered to have had a significant influence. If vaccinated people share positive messages with the unvaccinated, then vaccination could spread smoothly across the population. The effectiveness of the messaging by those who have been vaccinated is influenced by how strongly they recommend vaccination to others, and by their perceptions that led them to get vaccinated. These perceptions can be explained by the Health Belief Model (HBM) [11], which was developed as a disease conceptual model leading to health behaviors and has seven components, Perceived Susceptibility, Perceived Severity, Perceived Threat, Perceived Benefits, Perceived Barriers, Self-Efficacy, and Cues to Action. Recently. Several studies have reported the HBM-based analyses of factors influencing the decision-making to vaccinate against COVID-19 [12–14]. In addition, it is assumed that the degree of health literacy of the vaccinated person influences the nature of the messaging and vaccine recommendation to others [15]. However, few studies have reported on what messages were provided from vaccinated people based on their perspectives of the vaccination.

### 2.2. Objectives

In a qualitative study utilizing semi-structured interviews, our objectives were to understand the types of messages shared by healthcare providers and healthy adults receiving the COVID-19 vaccine to the unvaccinated people around them, and the relationship between the messages and the participant background, such as their perception and recommendation to the vaccine. Additionally, we set out to assess the differences and similarities between the healthcare providers and the healthy adults through these individual semi-structure interviews.

## 3. METHODS

### 3.1. Participants

A total of 10 participants including 5 healthcare providers and 5 healthy adults were targeted for recruitment in this exploratory study. The number of participants in this study was set at 10 since a previous qualitative study on vaccine hesitancy reported that data saturation was reached at around this number [16]. Three rounds of recruitment were pursued between August 23 and October 22, 2021 at the Chuo-city (Tokyo) administered vaccination site located at St. Luke’s Center for Clinical Academia, St. Luke’s International University. Participation in this study was voluntary and written informed consents were obtained.

Prospective participants who met the following criteria were considered for this study: 1) have completed the second dose of the COVID-19 vaccine, 2) healthcare providers, physicians or nurses, who were responsible for vaccination, or apparently healthy adults aged 20 years or older who were from the general population, 3) ability to communicate in Japanese, and 4) ability to conduct interviews via Zoom® (Zoom Video Communications, Inc., San Jose, CA, USA.). Exclusion criteria including the following: 1) persons with whom it was considered difficult to communicate directly, 2) those who were considered to have insufficient ability to understand and judge the interview, and 3) those who were judged to be difficult to conduct the interview with. These were assessed by the researcher during the screening.

After the participants were registered, the dates for the online interviews were set individually between August and November 2021. As the purpose of this research was to interview participants about their conversations with close persons after their vaccination, the online interview was scheduled to take place at least two weeks after the second vaccination.

The study was approved by the Research Ethics Board of St. Luke’s University, and given the approval number: 21-R076.

### 3.2. Questionnaire

After obtaining the participants’ consent, the questionnaire was used to collect information on socio-demographic characteristics and measures of health literacy. The questionnaire composed the following items: name, gender, age, date of birth, number of cohabiters, educational background, employment status, current health conditions, history of COVID-19 infection, the dates of the 1st and 2nd vaccinations, and the 14-item Health Literacy Scale for Japanese adults (HLS-14) [17]. The HLS-14 developed by Suka (2015) consists of three levels: “functional health literacy,” “communicative health literacy,” and “critical health literacy,” and has been widely used to assess the health literacy of Japanese adults [18–20].

### 3.3. Interviews

Interviews were practiced with an academic co-author and a medical colleague, beforehand to assess the appropriate interview time and the validity of the questioning items. The main interviews were conducted with each participant on different dates. Each interview took about 30-40 minutes and was recorded. A semi-structured interview consisting of 7 general, open-ended, neutral, and non-guided questions was prepared by the researcher and the co-author to evaluate the knowledge, attitude, behavior, and perceptions related to COVID-19 or COVID-19 vaccine. The interviews were conducted by the researcher with keeping in mind the components of the HBM. The HBM is a useful model for decision-making on vaccination [21–23]

#### 3.3.1. Semi-structured interview questions following the HBM

1. What do you know about the COVID-19 and COVID-19 vaccines? (Perceived Susceptibility, Perceived Severity, Perceived Threat)
2. What were your expectations about receiving the COVID-19 vaccine? (Perceived Benefit)
3. What were your concerns about receiving the COVID-19 vaccine? (Perceived Barrier)
4. Why did you decide to get the COVID-19 vaccine? (Cues to Action)
5. What is your opinion about whether to recommend the COVID-19 vaccine to unvaccinated people? (Vaccine Recommendation)
6. What did you talk about with your family, friends, and colleagues after you received the COVID-19 vaccine? (Messages after vaccination)
7. What is your general opinion about the COVID-19 vaccine?

### 3.4. Analysis

NVivo Qualitative Data Analysis Software Version 1.5.2 (QSR International, Massachusetts, U.S.) was used to analyze the qualitative data. The analysis procedure consists of the following steps: 1) Coding-identifying: in-vivo coding of comments relevant to the research in each interview; 2) Coding-sorting: sorting of duplicate comments in the code obtained in the first coding to create units; 3) Coding-condensation: enhancing the level of abstraction for units obtained in second coding; 4) Categorizing: creating categories by extracting from the third coding the content that matches HBM, vaccination recommendations, and the content that was talked about after vaccination; 5) Generalizing: generalizing multiple participant’s perspectives and significance albeit single-person perspectives based on the categories assigned to each item of the HBM. In order to eliminate arbitrariness as much as possible, each time one interview was completed, the review and analysis were carefully repeated by the researcher and the co-author.

## 4. RESULTS

### 4.1. Participants

Between August and October 2021, five healthcare providers who were engaged in vaccination and seven healthy adults who were vaccinated at the site were enrolled in the study (Fig 1). One of the healthy adults was not included due to scheduling difficulties and loss of contact. Individual interviews were conducted with a total of 11 participants, 5 with males and 6 with females (Table 1). Participant ages included two in their 20s, two in their 30s, five in their 40s, and two in their 50s. The mean period between the second vaccination and the interview was 114 days for health care providers and 21 days for healthy adults. Healthcare providers received the COVID-19 vaccine as a priority, therefore the mean period to interview was longer than with the healthy adults (Fig 2). The average time to conduct the interview was 35 minutes. The mean score of the HLS-14, a measure of health literacy, was 55.7±5.4 with no difference between healthcare providers (55.6±4.1) and the healthy adult population (55.7±6.1).

**Fig 1.**
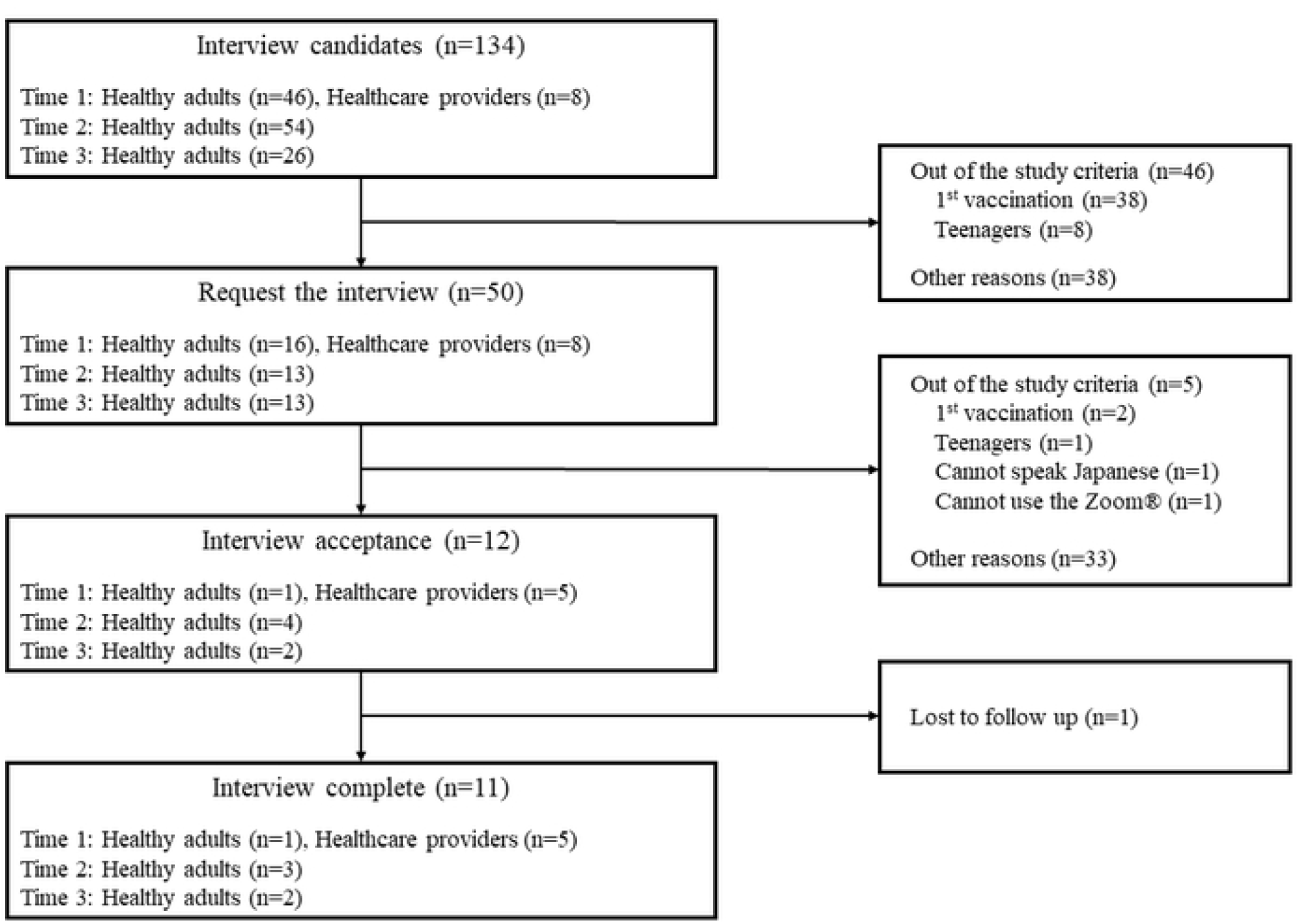
Flow chart of participant recruitment

**Fig 2.**
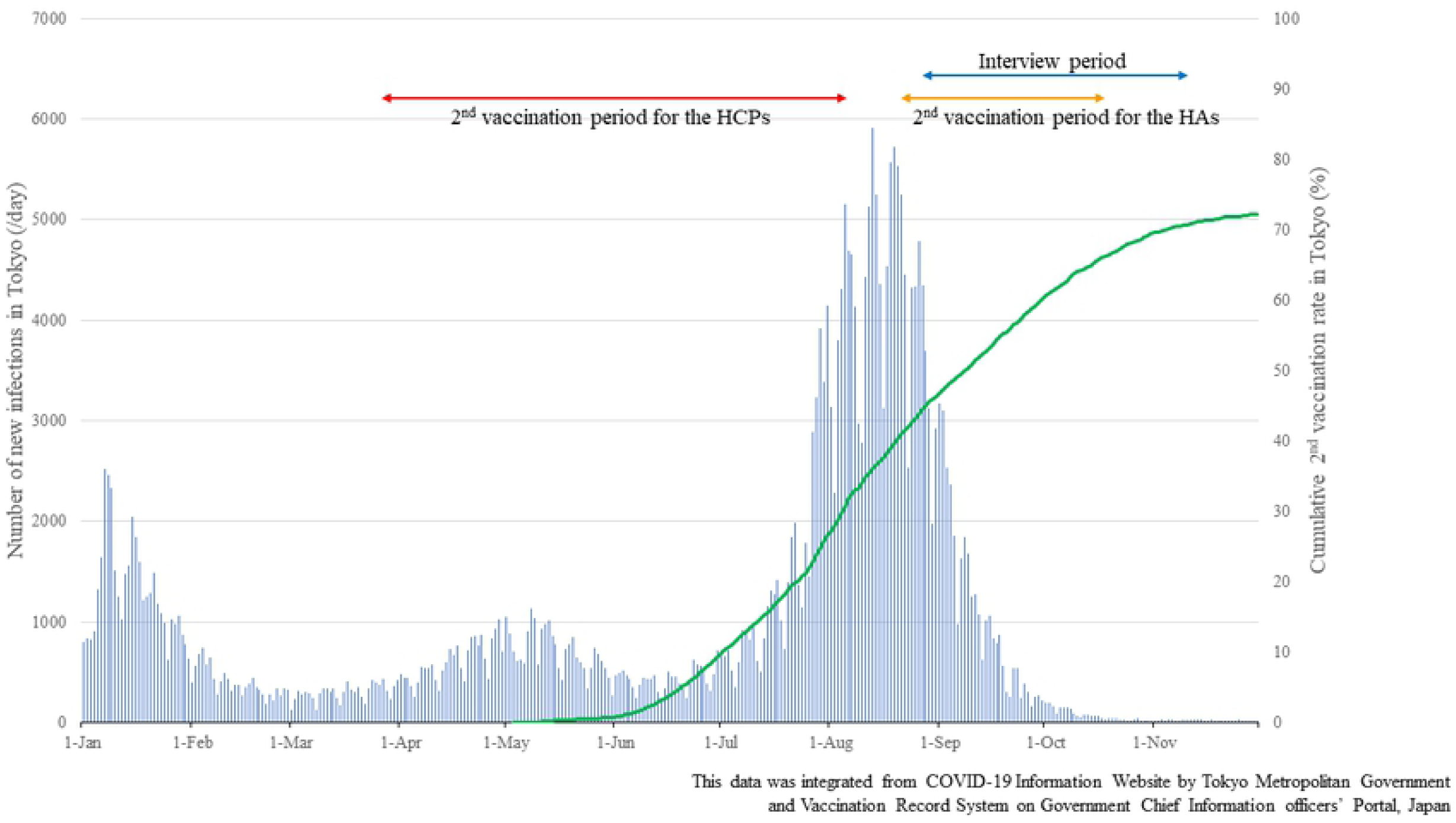
Number of the new infections and cumulative 2^nd^ vaccination rate in Tokyo, Jan.21-Nov.21 191

**Table 1.** Characteristic of participants

| No | Sex | Age('s) | HCP or HA | Number of Family Members Living with | Education | Employment Status | Infection history of COVID-19 |
| --- | --- | --- | --- | --- | --- | --- | --- |
| 1 | Male | 30 | Physician | 2 | University | Full time | No |
| 2 | Male | 40 | Physician | $\geq 3$ | Graduate school | Full time | No |
| 3 | Female | 50 | Nurse | 1 | Vocational school | Part time | No |
| 4 | Female | 30 | Nurse | 0 | Vocational school | Part time | No |
| 5 | Female | 40 | Nurse | $\geq 3$ | Junior college | Part time | Yes |
| 6 | Male | 40 | Healthy Adult | 1 | Graduate school | Full time | No |
| 7 | Female | 50 | Healthy Adult | 0 | Graduate school | Self-employed | No |
| 8 | Female | 50 | Healthy Adult | $\geq 3$ | University | Self-employed | Yes |
| 9 | Male | 40 | Healthy Adult | 1 | Graduate school | Full time | No |
| 10 | Male | 40 | Healthy Adult | 0 | Graduate school | Full time | No |
| 11 | Female | 20 | Healthy Adult | 0 | Vocational school | Full time | No |
| 12 | Female | 20 | Healthy Adult | 0 | University | Full time | No |

Table 1. Characteristic of participants continued
| No | Interview Month | Period between 2nd Vaccination and Interview | Duration of Interview (min) | HLS-14 Total Score |
| --- | --- | --- | --- | --- |
| 1 | Sep. 2021 | 5 months | 29 | 56 |
| 2 | Sep. 2021 | 5 months | 31 | 53 |
| 3 | Oct. 2021 | 5 months | 38 | 55 |
| 4 | Oct. 2021 | 2 months | 29 | 63 |
| 5 | Aug. 2021 | 2 months | 27 | 51 |
| 6 | Sep. 2021 | 20 days | 30 | 58 |
| 7 | Lost to follow up |  |  | 55 |
| 8 | Nov. 2021 | 1 month | 62 | 42 |
| 9 | Oct. 2021 | 22 days | 37 | 59 |
| 10 | Oct. 2021 | 15 days | 32 | 63 |
| 11 | Nov. 2021 | 24 days | 36 | 55 |
| 12 | Nov. 2021 | 16 days | 32 | 58 |

HLS-14: 14-item Health Literacy Scale for Japanese adults

### 4.2. Perceptions, Cues to Action, Vaccine Recommendation, and Messages

The results of 11 interviews included 117,040 words in Japanese spoken across 383 minutes of total interview time. The analysis for the interviews was carefully conducted with the co-author for about 10 hours. In total, 267 codes were identified among healthcare providers and 355 codes were identified among healthy adults, which were relevant to the purpose of the study (coding-identifying) (Table 2). By sorting the codes obtained from the coding-identifying step, 146 codes and 226 codes were identified (cording-sorting). By enhancing the level of abstraction for categorization, 94 codes and 140 codes were found, respectively (coding-condensation), and by categorizing the codes matching the HBM, 59 categories and 68 categories were created, respectively (categorizing). Finally, the generalizability of the categories was 26 and 27 in healthcare providers and healthy adults, respectively (generalizing). The categories for vaccination recommendations were 3 and 6, and the categories for messages after vaccination were 3 and 7 for healthcare providers and healthy adults, respectively. Each category and main comments are listed (S1 and S2 Table). and S2 Table. Also, summary of the results is indicated (Fig 3).

**Table 2.** Analytical process for the interviews

|  | **Participants** | **1<sup>st</sup> Coding:**<br>**Identifying** | **2<sup>nd</sup> Coding:**<br>**Sorting** | **3<sup>rd</sup> Coding:**<br>**Condensation** | **Categorization** | **Generalization**<br>**of Perceptions** | **Vaccine**<br>**Recommendation** | **Messages** |
| --- | --- | --- | --- | --- | --- | --- | --- | --- |
| HCP | 5 | 267 | 146 | 94 | 59 | 26 | 3 | 3 |
| HA | 6 | 355 | 226 | 140 | 68 | 27 | 6 | 7 |

HCP: Healthcare Provider, HA: Healthy Adult

**Fig 3.**
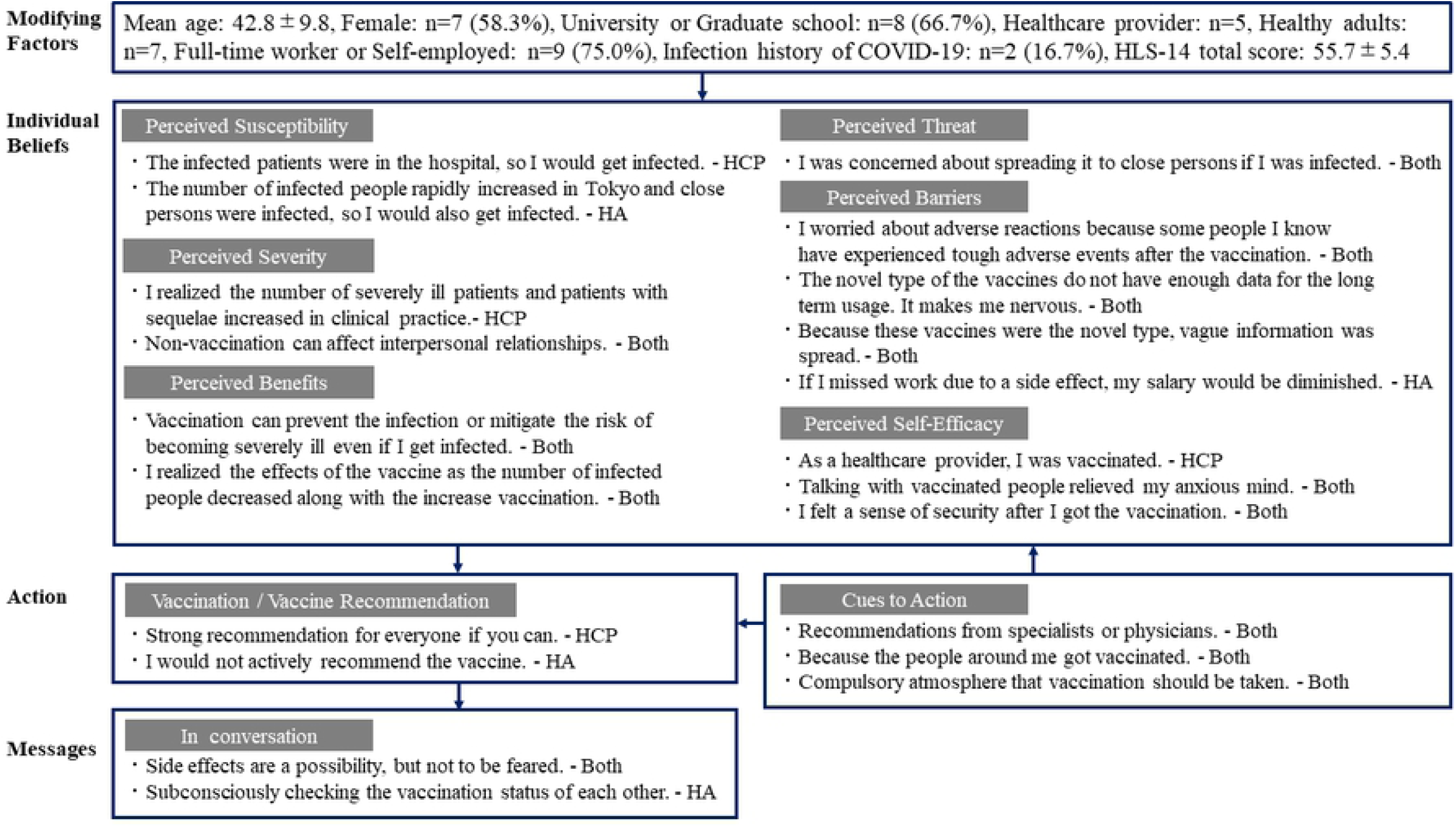
Summary results based on the Health Belief Model

#### 4.2.1. Perceived susceptibility

The healthcare providers were aware of the possibility that they might be infected with COVID-19 at their hospital, even if they were not directly involved in the treatment because the hospital where they worked treated COVID-19 infected patients. On the other hand, the healthy adults began to be aware that they might also be infected when the number of infections rapidly expanded in Tokyo (Fig 2) [24], or when they saw or heard that there were infected people nearby. However, when there were no infected people in their immediate vicinity, the sense of reality was weakened, leading them to think that they might not be infected weakening their perceived susceptibility.

#### 4.2.2. Perceived Severity

The healthcare providers realized that the severe cases of COVID-19 increased in their daily practice and that most of the severe cases were unvaccinated patients. Furthermore, even when discharged from the hospital, some patients still suffered from sequelae, and this made them aware of the COVID-19 threat.

In addition, some participants realized that not getting vaccinated may affect their interpersonal relationships; for example, by wearing a mask when meeting unvaccinated people. The healthy adult #12 has had similar experiences about relationships with others and realized that the conversation atmosphere deteriorated when she mentioned to others that she had no intention to vaccinate. The healthy adults were concerned that not vaccinating may lead to discrimination and prejudice. On the other hand, the healthy adult #8 commented that since there were more people infected with the COVID-19 in Tokyo than in rural areas, she did not face discrimination or prejudice from others even when she was infected. It is possible that whether the number of infected people is high or low in the surrounding area may influence the tendencies for discrimination and prejudice.

#### 4.2.3. Perceived threat

Both the healthcare providers and the healthy adults were threatened by their own infection with the COVID-19 which could lead to secondary infection of those around them. In particular, those who lived with their families were more aware of the need for vaccination to protect family members.

#### 4.2.4. Perceived benefits

The perceived benefit was similar for both the healthcare providers and the healthy adults. They hope that vaccination will suppress the transmission of the COVID-19 and reduce the risk of getting severely ill if infected. At the time of the interviews, the number of infected and severe cases in Tokyo were decreasing, making them realize the effect of the vaccine because the vaccination had gradually spread (Fig 2) [25]. They expected to be able to have dinner with friends, travel, and return to their hometowns, if the number of infected and severe cases continued to remain low. Such expectations were clarified by the interviews.

#### 4.2.5. Perceived barriers

Many comments were received related to the perceived barriers, both from the healthcare providers and the healthy adults. The most common perception was concern about side effects from the vaccination. People generally imaged the vaccines to resemble influenza vaccines, and were not aware that influenza vaccination can cause side effects. However, since the CIOVID-19 vaccine has a higher rate of side effects than conventional vaccines [26], they realized that it was completely different experience. They felt even more anxious before their vaccination after hearing about the painful side effects from the vaccinated people.

Also, the COVID-19 vaccines were a novel type with a different development process than the conventional ones, and people felt anxious about the novelty itself. Because of the lack of sufficient long term clinical data, they were concerned about unexpected side effects and sequelae in the future. As it was a novel type of vaccine, there was a lot of uncertain information on social networking sites and the Internet, such as “the COVID-19 vaccine will lead to infertility” and “the COVID-19 vaccine will affect immunity and eventually cause death”. These questionable information had led to a sense of barrier to vaccination.

The unique perception of the healthy adults was that they recognized that Spikevax™ causes more side effects than Comirnaty™. While the healthcare providers did not have the option to vaccinate other than Comirnaty™, the healthy adults could choose amongst Comirnaty™, Spikevax™, or Vaxzevria™, causing a sense of barrier due to the difference in safety between the vaccines.

Employment status was also thought to have an impact on the sense of barrier to the vaccination. Since many young people, in particular, were non-regular workers [27], they were concerned about the influence of side effects of the vaccine on their work. Since there was no compensation for their salary if they missed work due to side effects, they were worried about decreased income. Health adult #12 had heard such opinions from her close persons.

Furthermore, healthcare providers’ vaccination schedule was set by the hospital where they work, so they did not need to make their reservations for vaccination. However, the healthy adults can decide to get vaccinated at their own timing. Therefore, some healthy adults were aware that they did not need to be vaccinated immediately. This could be a factor that enhances the sense of barrier. On the other hand, the majority of the participants felt that side effects after vaccination were mild and not as painful as they had expected, and this was not a barrier to recommending vaccination to others.

The participants recognized the importance of providing correct information, as they sometimes saw or heard ambiguous information. The healthcare providers were aware from their vaccination work that many people had concerns about vaccines, and believed that it was necessary to have a point of contact for these people to feel free to talk about various concerns and worries about the COVID-19 vaccines.

#### 4.2.6. Self-efficacy

The healthcare providers had made the decision to vaccinate because of their sense of responsibility. As healthcare providers, they tried to give correct information to their family and friends, as they were often asked about vaccinations. To enhance self-efficacy, they believed it is important to know about the COVID-19 vaccines first.

Both the healthcare providers and the healthy adults found that their concerns before vaccination were alleviated by talking with others who had been vaccinated, and they felt more secure after vaccination. On the other hand, those who did not originally intend to be vaccinated did not have a high sense of self-efficacy even after receiving the vaccine.

#### 4.2.7. Cues to Action

Both the healthcare providers and the healthy adults were motivated to get vaccinated by recommendations from specialists of infection disease and physicians, and by the fact that the people around them had also been vaccinated.

Even though they were not forced to be vaccinated, they felt the atmosphere in their workplace that they should be vaccinated. The ease of making reservations for vaccination was also a trigger for vaccination, as the healthy adults had to make their own reservations.

#### 4.2.8. Vaccine recommendation to others

The healthcare providers were very eager to have as many people as possible vaccinated. As the COVID-19 vaccines were the most promising way to reduce the risk of getting severe infection, and realizing how devastating it can be, they believed that vaccines have more benefits than risks.

On the other hand, recommendations from the healthy adults for vaccination of others were varied. Some recommended vaccination to close family members, but did not actively recommend vaccination to others. This is because they recognize that not getting the vaccine is an option and should not be forced. Therefore, they would like to respect the unvaccinated people’s opinions. Differences in vaccine recommendations were found between the healthcare providers and the healthy adults.

#### 4.2.9. Messages

Both the healthcare providers and the healthy adults often talked about side effects with their family, friends, and colleagues after vaccination. However, they did not try to stir up concerns, but rather told others that based on their own experiences the vaccination did not need to be scary and that they felt relieved after the vaccination. This was because the participants in this study did not experience any strong side effects from the vaccination, and that the vaccination reduced the risk of infection and becoming severely ill from the COVID-19, as well as the risk of transmission to others.

A feature of the healthy adults was that they unconsciously checked the other person’s vaccination status during the conversation. The vaccination status was a common topic of conversation in the COVID-19 era, and people naturally shared their vaccination experiences. This shared experience eased the fear of the COVID-19 vaccine for those who had not been vaccinated. By contrast, those who had no intention to be vaccinated felt stressed by having their vaccination status checked and by being implicitly recommended or pressured to be vaccinated.

## 5. DISCUSSION

This study was the first to understand what messages were shared by the vaccinated healthcare providers and healthy adults after their vaccination, and the relation of the individual’s thoughts and social background in influencing the messages. Both of themshared similar messages from their own vaccination experiences to ease others’ concerns about side effects of the COVID-19 vaccination. However, differences in vaccine recommendations were observed between the healthcare providers and the healthy adults. The mean score of the HLS-14 did not differ between the two groups. Therefore, it was not clear whether health literacy affected the contents regarding what they talked about after vaccination. We described here the backgrounds and relationships that influence vaccine recommendations in both the healthcare providers and the healthy adults.

### 5.1. Vaccine recommendations from the healthcare providers

The healthcare providers who participated in this interview were unanimous in their recommendation for vaccination. The strongest reason for this is that they expect that vaccination will prevent getting severely ill even if infected with COVID-19. In their daily clinical practice, the healthcare providers have treated patients suffering from different kinds of diseases and have seen patients with significant functional disabilities, severe diseases, and die from diseases. Therefore, it was believed that they were treating patients with a strong motive to keep them from becoming severely ill and reduce deaths caused by diseases as much as possible. According to the perceived severity, we found that the healthcare providers strongly recognized the severity of COVID-19, such as the fact that unvaccinated patients with COVID-19 infection could become severely ill, the sequelae of COVID-19 infection could persist, and the sense that the number of the severe cases was increasing. Vaccination has been recognized as a useful measure to reduce the risk of getting severely ill. The reason why they recommend vaccination was not only because of its advantage, but also because of the downside of not being vaccinated which would change the hospital’s acceptance of an infected patient who becomes severely ill. Of course, they were concerned about side effects of the vaccine and the lack of long-term clinical data. However, based on the current data and their experiences and those of their colleagues, few serious side effects occurred, and they believed that the benefits of vaccination outweigh the risks at this point in time.

The interviews also clarified that the healthcare providers felt strongly about their responsibility. In the initial phase of the vaccination in Japan, when the experience of COVID-19 vaccination was still minimal, their sense of responsibility as healthcare providers promoted vaccination. And in the conversation with their close persons after the vaccination, healthcare providers mainly talked about side effects based on their own experiences, but they also emphasized that there was no need to be afraid of side effects. This was because many people were worried about side effects of the vaccines, so they were trying to alleviate the concerns by explaining that side effects were milder than they had expected. In addition, they told the others that the vaccination gave them a sense of security because it reduced the risk of getting severe illness even if infected with the COVID-19.

### 5.2. Vaccine recommendations from the healthy adults

The level of vaccine recommendations from the healthy adults was inconsistent. While some healthy adults were willing to recommend vaccination to close family members, others were not actively recommending it to others. This may be affected by how much expectation they had for the vaccine in advance. Those who recommended vaccination were more likely to have been living with family members, and as a perceived threat, they were worried about transmitting COVID-19 to others if they had been infected. The recommendation to vaccinate might have been based on their desire to protect the family. In contrast, those who responded that they would not actively recommend the vaccine did not live with their families, but were considering getting the vaccine when the number of infected people significantly increases in Tokyo (Fig 2). This tendency was observed among younger people who, besides being concerned about side effects from the vaccine, were also concerned about the fact that it was a novel type of vaccine. Especially for non-regular employees, they were worried about loss of income if they had to take a leave due to side effects of the vaccine, which suggests that their expectations of the vaccine were relatively low. Not only the expectation and insecurity toward the vaccine, but also environmental factors such as living with family members and employment status may affect the level of recommendation after vaccination.

In conversations after vaccination, the healthy adults mainly talked about side effects, based on their own experiences, to close persons and people who were worried about getting vaccinated, but told them that there was no need to worry. The message itself is the same for the healthcare providers and the healthy adults. A feature of conversations among the healthy adults was that they unintentionally checked each other’s vaccination status during the conversation. This may be indicating an unconscious interest in recognizing the risk of infection that affects them. If the other person in the conversation was unvaccinated, they might refrain from future contact or hope that the other person would be vaccinated as well. On the other hand, unvaccinated people would feel that being checked about their vaccination status or being recommended for vaccination itself was vaccine-related harassment. This point should be taken into consideration when recommending vaccination.

### 5.3. Suggestions for practice

Lastly, the suggestions for practice in order to further promote vaccination are discussed from a public health perspective. Removing the concerns about the vaccine may lead to vaccination, which in turn may influence vaccine recommendations to others. In the interviews, it was found that anxiety about vaccines can be alleviated by sharing the experience of vaccination, that some unvaccinated people are not able to ask about their vaccine anxiety even if they want to, and that some people are reluctant to get vaccinated because they are concerned about their income loss due to side effects. To address these issues, we suggest the following measures into the practice.

#### 5.3.1. Share the experience with people close to you after vaccination

It has been reported that information from healthcare providers and people close to the target patients have more influence on health behavior than information from the media or the Internet [7]. And those who were hesitant to get vaccinated but did get vaccinated trusted information from healthcare providers and people close to them, such as family members[6]. By sharing the experience after vaccination, the other person who hears about it can gain a sense of security. By making their vague concerns concrete, those who are vaccinated may be able to ease the distrust that unvaccinated people may have towards the vaccines.

However, in sharing experiences, we need to be careful in our conversations to avoid unconsciously checking the other person’s vaccination status and implicitly coercing them into vaccination. This is because people who have decided not to vaccinate feel stressed by having their status checked and being implicitly coerced to get vaccinated.

#### 5.3.2. Set up a point of contact at vaccination sites where people can feel free to ask about vaccines

People who have been vaccinated, those who are planning to be vaccinated, and those who have no intention to be vaccinated have concerns. They are concerned about the safety of vaccines, including the long-term effects, the fact that each vaccine causes different side effects, and the risk of infection or becoming severely ill if they are not vaccinated. First of all, knowing exactly what the vaccine is may relieve some of their concerns, and talking about it can give them a sense of security. If they are still feeling insecure toward vaccination, positive messages will not be shared after vaccination. Eliminating concerns before vaccination is important in the dissemination of messages after vaccination.

#### 5.3.3. Compensation for non-regular employees

In Japan, the proportion of non-regular employees is high, especially among women and young people [27]. Household income has been reported to be a factor in vaccine hesitancy [28]. Non-regular employees are concerned not only about side effects of the vaccine, but also about the possible loss of income due to the difficulty of working as a result of side effects. They have limited paid holidays compared with regular employees, and leave due to side effects may be counted as absenteeism. Therefore, if non-regular employees are vaccinated and have to be absent at work due to side effects, compensating them financially could help promote vaccination.

### 5.4. Limitations

There were several possible limitations in this study. At the vaccination sites in Chuo City, only residents of the Chuo City were eligible for vaccination. The characteristics of the participants include a high percentage of those with a university or graduate school education and a high level of health literacy. According to the national census in Japan [29], the national average of those with a university or graduate school education was 19.9%, while the participants in this study was 66.7%. The mean HLS-14 score in the national survey conducted by Suga (2013) was 50.3[17], while that of the participants in this study was 55.7, suggesting the possibility of selection bias and a higher level of health awareness than people in general. Therefore, most participants have had the thought of recommending vaccination.

Furthermore, since only Comirnaty™ was provided at the vaccination site in this study, comments from people who had taken other vaccines were not available. Although the side effects reported for each vaccine were different, the incidence rate of side effects did not differ significantly for any COVID-19 vaccines, so the influence on the results of this study was considered to be minimal[26].

Elderly people and those under 20 years of age were not enrolled in this study. Enrollment in the study did not begin until August 2021, and many elderly people had already been vaccinated, so it was difficult to recruit them. Those under 20 years of age were also excluded because of the possible influence of their parents on their vaccination decisions [30,31]. If elderly people or young people participate in this study, it may affect the results of the vaccine recommendation and the messages. Further studies may be needed for these populations and the unvaccinated people.

### 5.5. Conclusions

In this study, the messages from the conversations between the participants and those around them after vaccination were examined, as well as the relationship between the messages and the individual’s thoughts and social background based on the HBM. Both the healthcare providers and healthy adults shared similar messages to ease the vaccination concerns of others regarding side effects. However, their vaccine recommendation level was varied, which may be influenced not only by expectations and concerns toward the vaccine, but also by external factors such as family members living together.

## Data Availability

The data for this study consisted of transcripts of 11 participants, including identifying information even though they are anonymized. This data cannot be shared to the public due to participant confidentiality and ethical requirements. Participants consent to this study under the condition that their anonymity is ensured. Therefore, only illustrative quotes from the transcripts are included in this paper. Data access requests may be made to St. Luke's International University at Institutional Review Board.

https://stopcovid19.metro.tokyo.lg.jp/

https://cio.go.jp/vrs

## 7. Supporting information

S1 Table. Healthcare Providers’ perceptions for each category of the Health Belief Model

S2 Table. Healthy adults’ perceptions for each category of the Health Belief Model

